# Feasibility study of the application of Magnetic Resonance Elastography (MRE) to diagnose adenomyosis

**DOI:** 10.1101/2024.09.03.24313024

**Authors:** V Jain, E Hojo, G McKillop, A Oniscu, Y Le, J Chen, R Ehman, N Roberts, HOD Critchley

**Affiliations:** Centre for Reproductive Health, Institute for Regeneration and Repair, University of Edinburgh, Edinburgh, United Kingdom; Royal Infirmary Edinburgh, Edinburgh, United Kingdom; Mayo Clinic, Rochester, Minnesota, USA; Department of Radiology, Nanjing Drum Tower Hospital, Nanjing, China; Resoundant, Inc., Rochester, MN, USA

**Author notes:** Joint Corresponding Authorship: Professor Hilary Critchley and Professor Neil Roberts: Centre for Reproductive Health, Institute for Regeneration and Repair, University of Edinburgh, 4-5 Little France Drive, Edinburgh, EH15 4UU. and. E. Hojo - Mayo Clinic, Rochester, Minnesota, USA. A. Oniscu - Karolinska University Hospital, Sweden.

**Keywords:** adenomyosis, magnetic resonance elastography (MRE), diagnosis, fibrosis, abnormal uterine bleeding

## Abstract

**Introduction:** Adenomyosis is an under-recognised condition in which definitive diagnosis is only possible via histology after hysterectomy, an unacceptable option for those wishing to preserve fertility. Recent cellular/molecular studies indicate adenomyotic lesions may be fibrotic leading to increased uterine tissue stiffness. 3D Magnetic Resonance Elastography (MRE) is a novel imaging technique that allows in vivo measurement of tissue stiffness (via elastograms). 3D MRE has not been reported to study adenomyosis. The feasibility study aimed to utilise a novel 3D MRE protocol to measure global uterine stiffness and to investigate its potential application for non-invasive in vivo diagnosis of adenomyosis.

**Materials and Methods:** 3D MRE protocol was conducted on one healthy volunteer (control) and four patients with suspected adenomyosis and heavy menstrual bleeding (HMB), diagnosed via transvaginal ultrasound and clinical history (REC:20/SS/0123 and 19/SS/0102). Two patients underwent hysterectomy, and representative uterine tissue samples were assessed for (i) histological presence of adenomyosis via H&E staining; (ii) cellular/molecular measures of tissue stiffness (collagen [picrosirius red], α-smooth muscle actin, e-cadherin); (iii) relationship between in vivo assessment of the uterus via MRI images and 3D MRE findings with in vitro uterine tissue histology from the same individuals.

**Results:** 3D MRE was successfully used to acquire elastograms for four patients with adenomyosis (diffuse n=3, focal n=1) and one healthy volunteer. Calculated global uterine stiffness was higher in women with adenomyosis (2.93kPa; range 2.34 – 3.39kPa) compared to a healthy volunteer (2.04kPa). Areas of stiffness on 3D elastograms reflected adenomyotic changes visualised via conventional MRI, with the added benefit of also correlating with histology/immunohistochemical assessment for markers of tissue stiffness.

**Discussion:** A novel 3D MRE protocol has been applied to obtain the global uterine stiffness in four women with HMB and suspected adenomyosis, and one healthy volunteer. 3D MRE has the potential to provide superior non-invasive tissue characterisation in vivo when compared to conventional MRI in the assessment of adenomyosis due to the correlation of imaging and tissue findings. Further studies are now needed to confirm the above exploratory findings, prior to performing a potential clinical trial.

## Introduction

Adenomyosis is a debilitating uterine disorder that is associated with heavy menstrual bleeding (HMB) and painful periods. It is defined as the presence of endometrial glands and stroma in the myometrium at least 2.5 mm from the endo-myometrial junction (Bird, McElin, and Manalo-Estrella 1972; Camboni and Marbaix 2021). Definitive diagnosis of adenomyosis can be obtained by a hysterectomy however this is a major, invasive gynaecological operation that is also fertility-ending, and not acceptable to many reproductive-aged women. Presently, there is a lack of an accurate, reproducible, non-invasive diagnostic modality that can equal the histological definition and as a consequence, the condition remains under-diagnosed and under-researched. Transvaginal UltraSound (US) and Magnetic Resonance Imaging (MRI), have been investigated as potential non-invasive techniques for diagnosing suspected adenomyosis. The reported sensitivities of 83%, and 88%, respectively (Meredith, Sanchez-Ramos, and Kaunitz 2009; Novellas et al. 2011) are reasonable but not always achieved in practice. In this study, we tested the feasibility of employing an MRI modality known as Magnetic Resonance Elastography (MRE) for diagnosing adenomyosis.

Due in part to the above diagnostic challenges, there is currently a lack of understanding regarding the aetiology and pathophysiology of adenomyosis (Guo 2022). One theory is that after an area of adenomyosis (ectopic endometrial epithelial glands and stoma) is established within the myometrium, it may undergo cyclical changes in response to fluctuating levels of circulating sex steroid hormones, and similar to the eutopic endometrium, the area of adenomyosis may bleed. (Leyendecker, Wildt, and Mall 2009; Vannuccini et al. 2017). Subsequently, adenomyosis progresses to a state where it is characterised by fibrosis which occurs through epithelial–mesenchymal transition (EMT), fibroblast-to-myofibroblast trans-differentiation (FMT) and smooth muscle metaplasia (SMM) (Liu et al. 2016; Huang et al. 2022)

MRE allows non-invasive quantification of the mechanical properties of living tissues (Manduca et al. 2021) and, given the above-mentioned significance of the occurrence of fibrosis, the technique is potentially well suited to supporting the diagnosis of adenomyosis. MRE protocols are already well established and clinically validated for staging the severity of liver fibrosis (Ehman 2022). There have also been reports of the application of 2D MRE in the study of the uterus (Stewart et al. 2011; Jondal et al. 2018; Obrzut et al. 2020) and which demonstrated that 2D MRE was successful in detecting and characterising leiomyomas but adenomyosis was not studied.

The primary objective of the present study was to apply a novel protocol utilising state-of-the-art 3D MRE to measure global uterine stiffness in four patients with suspected adenomyosis. To our knowledge, this is the first study where 3D MRE of the in vivo uterus with adenomyosis, has been accompanied with the histological assessment of the same uterus in vitro. Therefore, the secondary objective was, in two patients who went on to have a hysterectomy shortly after having 3D MRE scans, to correlate imaging findings with histological investigations of the uterus, including markers of stiffness, to determine if 3D MRE is consistent with histopathology. We hypothesised that 3D MRE has potential to non-invasively diagnose adenomyosis, with correlation between T2 weighted MRI findings, histological findings from matched stained uterine tissue sections, and an increased tissue stiffness on the 3D MRE elastograms.

## Materials and Methods

### Patient recruitment

The study received respective approvals from the NHS Health Research Authority’s Research Ethics Committees which permitted the recruitment of healthy volunteers (control) to support the development of imaging protocols testing (Ref: 20/SS/0123), together with patient recruitment (study participant) and use of their uterine tissue samples (Refs. 19/SS/0102 and 20/ES/0119). Fully informed written consent was obtained from all study participants, with one control recruited from the University of Edinburgh, and four study participants recruited after attending the gynaecology outpatient services at the Royal Infirmary Edinburgh, NHS Lothian. At recruitment, the control, who was of reproductive age, did not report any clinical history of gynaecological complaints and had a normal menstrual cycle (as per International Federation of Obstetrics and Gynecology (FIGO) AUB system 1) (Munro et al. 2011; Munro et al. 2018). None of the participants were using hormonal treatments at the time the study was conducted. All four study participants were women of reproductive age, with the complaint of HMB and suspicion of adenomyosis following transvaginal US which was reviewed in accordance with Morphological Uterus Sonographic Assessment (MUSA) guidance on ultrasound diagnosis of adenomyosis (Van den Bosch et al. 2019; Harmsen et al. 2022). There was no evidence of uterine fibroids being present in any of the participants.

A flow diagram of the study is shown in Figure 1. The same MR investigation, comprising T2-weighted MRI and 3D MRE imaging was performed for one control and four study participants, for two of whom a hysterectomy was performed for definitive treatment of HMB within 2 days following the MRI and 3D MRE imaging.

**Figure 1.**
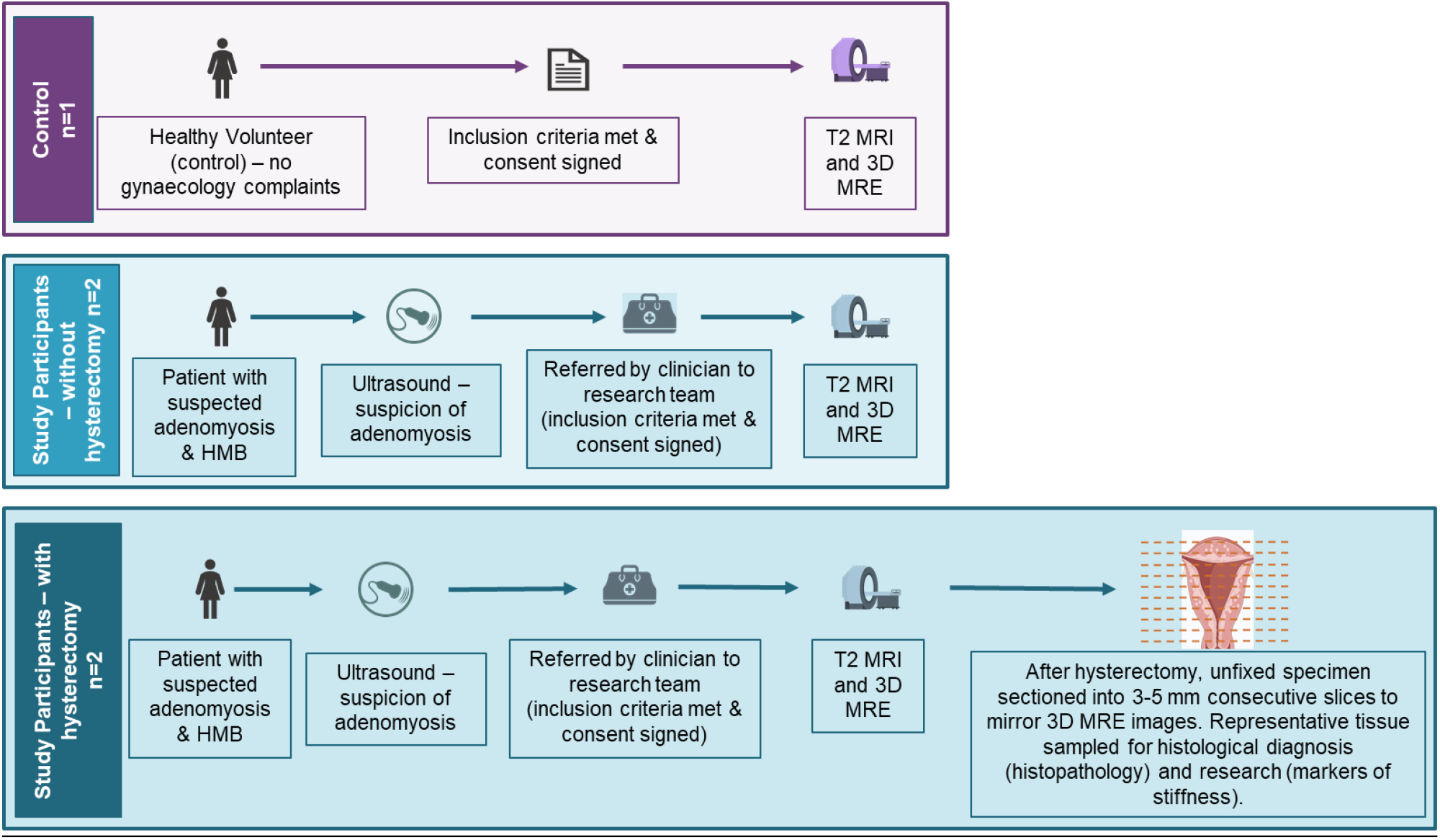
The pathway followed by one healthy volunteer (control) and the study participants who did and did not have hysterectomy. Study participants were patients with suspected adenomyosis (based on transvaginal ultrasound scan and clinical history) and heavy menstrual bleeding (HMB). Hysterectomy occurred after the T2 weighted MRI and 3D MRE imaging. Hysterectomy specimens (unfixed) were sectioned to mirror the planes from the 3D MRE images. MRI = magnetic resonance imaging, 3D MRE = three dimensional magnetic resonance elastography. [to the reproduced in colour in print and on the web]

### Setup for Acquisition of MRI and MRE Data

MR investigations were performed at the Edinburgh Imaging Facility at The Queen’s Medical Research Institute (EIF-QMRI), University of Edinburgh using a 3 T Skyra Fit MRI system (Siemens Healthineers, Erlangen, Germany). The participant lay supine and a 32-channel spine coil was positioned posteriorly and a 30-channel body matrix coil anteriorly. The MRE actuator, which has previously been used to perform cardiac MRE (Arani et al. 2017), was placed anteriorly on the lower abdomen, above the pubis symphysis and medial to the anterior superior iliac spine regions to avoid close proximity to bony structures. A foam cube was placed between the actuator and the body to provide good coupling of the acoustic waves. Both actuator and foam were secured by using an elastic belt which ensured good contact between the actuator and the body and maintenance of a central position. The actuator was connected via a plastic tube allowing pneumatic transmission of vibrations from the active driver of the Resoundant MRE system (Mayo Clinic, Minnesota, USA) to the MRE actuator (i.e., passive driver).

### MR Image Analysis

For each participant co-aligned and co-localised series of T2 weighted anatomical images and 3D Echo Planar Imaging (EPI) MRE images were acquired in identical true axial orientation, with Field of View (FOV) 24 cm and slice thickness 3 mm covering the whole uterus. For the T2 weighted MR images, the imaging matrix was 640 pixels by 640 pixels, TR 7570 msecs and TE 86 msecs and the acquisition time was 4 minutes 40 seconds. For MRE, a vibration frequency of 60 Hz was chosen, and the imaging matrix was 80 pixels by 80 pixels, TR 6400 msecs and TE 79 msecs. For each axial level a stiffness map was produced by using a 3D wave inversion algorithm and output directly on the MRI system. The acquisition time was 5 minutes 33 seconds.

### MR Image Analysis

The T2-weighted images were reviewed by a Radiologist (GMcK) in consideration of whether adenomyosis could be confirmed. MRE data were analysed by using ITK-SNAP (Yushkevich et al. 2006). In particular, a Region of Interest (ROI) was drawn demarcating the whole uterus as it appeared on the T2-weighted image and transferred and superimposed on the relevant stiffness map. The mean stiffness (kPa) value for the image pixels corresponding to the selected ROI and its variance were computed. This procedure was repeated for all the section levels encompassing the whole uterus. Significance testing did not occur due to the sample size of this feasibility study.

### Tissue Preparation and Sampling

Following hysterectomy, the fresh unfixed uterus was serially sliced into transverse sections using as a guide the approximate locations of the imaging planes that were prescribed in the MR investigation. Biopsies were taken so as to sample myometrial and endometrial tissue in anterior, posterior, right lateral, left lateral and fundal regions of the uterus. A sample was also obtained of endometrial tissue in the uterine cavity by using a Pipelle® endometrial suction curette (Pipelle de Cornier Mark II, Laboratoire CCD, France).

### Tissue Processing and Staining

Uterine tissue samples were fixed in 4% neutral buffered formalin and embedded in paraffin using standard procedures. Tissue sections with 4µm thickness were cut from the samples and stained with haematoxylin and eosin (H&E) and picrosirius red (0.5% Direct Red (Sigma-Aldrich) in Picric acid (Sigma-Aldrich)). In addition, immunohistochemistry assays were performed automatically for e-cadherin (Cell Signalling Technologies, a marker of EMT) and alpha smooth muscle actin (Sigma-Aldrich, αSMA; a marker of FMT) monoclonal antibodies by using the Leica Bond-Max autostainer (Leica Microsystems GmbH, Wetzlar, Germany).

Histopathologist (AO), who was blinded to the sampling location, reviewed the H&E stained tissue sections to potentially confirm a diagnosis of adenomyosis, and the picrosirius red, αSMA and e-cadherin stained slides to confirm the presence or absence of collagen fibres, smooth muscle fibres and epithelial cells, respectively.

### Correlation of MR Imaging and Uterine Tissue Staining

Once pathological reporting had been completed for the uterine tissue samples and MR images, and stiffness measurements had been obtained, a Radiologist (GMcK), Histopathologist (AO) and Gynaecologist (VJ) systematically reviewed the T2-weighted images for each of the four patients, as well as the stained tissue sections, for the two patients who underwent hysterectomy. The stained uterine tissue sections and corresponding MR investigations were reviewed together to confirm if the histologically confirmed areas of adenomyosis were detectable on the T2 weighted MRI. All reviewers were blinded to the findings of the 3D MRE investigations at this stage. The regions of the confirmed adenomyosis were subsequently mapped to the 3D MRE elastograms to correlate whether they displayed increased tissue stiffness.

## Results

All participants completed the study. No participant was menstruating at the time the study was performed. No suspected adenomyosis was observed on the MR images obtained for the control subject. For all four study participants who had been diagnosed as having suspected adenomyosis on the basis of transvaginal ultrasound performed prior to their participation in the study, the MR images acquired during the study confirmed the same diagnosis. Furthermore (see Table 1), the systematic review of the T2-weighted MRI images, stained uterine tissue sections, and 3D MRE elastograms (stiffness maps) obtained for the two study participants in whom a hysterectomy was performed, provided qualitative support for the prediction that regions which the Radiologist diagnosed as adenomyosis, corresponded to regions where the Histopathologist confirmed there is evidence of fibrosis, together with increased stiffness on the 3D MRE elastograms.

**Table 1.** Summary of the transvaginal ultrasound, Magnetic Resonance Imaging (MRI), 3D MRE and uterine tissue staining observations for participants, as applicable per study participation. * after study completion, this patient (Ad4) underwent hysterectomy for treatment of heavy menstrual bleeding, and received a histologically confirmed diagnosis of diffuse adenomyosis with probable adenomyoma.

| Participant | Transvaginal ultrasound | T2 MRI | Histopathology | Markers of tissue stiffness |  |  |  |
| --- | --- | --- | --- | --- | --- | --- | --- |
| | | | | 3D MRE (marker of tissue stiffness with imaging) | Picrosirius Red staining | Alpha smooth muscle actin (αSMA) staining | E-cadherin staining |
| **Control** | No ultrasound performed | No adenomyosis suspected in the uterus | N/A | Elastograms showed low uterine stiffness |  |  |  |
| **Study participant Ad1** | Heterogenous appearances of the uterus with an ill-defined mass 26 x 22 x 25 mm, ?fibroid, however the anterior wall of the uterus is thicker than the posterior wall | Diffuse and homogenous junctional zone expansion with only trivial submucous cystic change consistent with diffuse adenomyosis | Confirmatory regions of adenomyosis detected throughout the uterus, no evidence of fibroids. | Areas of increased stiffness throughout uterus | Increased collagen deposition throughout uterus except in regions of adenomyosis at sites of ectopic epithelial glands and stoma | αSMA staining throughout myometrium – less in regions of adenomyosis | e-cadherin staining identified in epithelial glands of the regions of adenomyosis confirming presence |
| **Study participant Ad2** | Heterogenous myometrium suggestive of adenomyosis | Junctional zone irregularly thickened with focal areas of cystic change in keeping with significant and extensive adenomyosis | Confirmatory regions of adenomyosis detected throughout the uterus | Areas of increased stiffness throughout uterus | Increased collagen deposition throughout uterus except in regions of adenomyosis at sites of ectopic epithelial glands and stoma | αSMA staining throughout myometrium – less in regions of adenomyosis | e-cadherin identified in epithelial glands of regions of adenomyosis confirming presence |
| **Study participant Ad3** | Uterus is bulky with increased echogenicity in the anterior wall and some cystic change with increased vascularity suggestive of adenomyosis | Possibly very early focal thickening of the junctional zone suggesting focal adenomyosis but not sufficient to call clinical diagnosis of adenomyosis | N/A | Targeted area of increased stiffness in the uterus to correlate with region of focal adenomyosis seen on MRI |  |  |  |
| **Study participant Ad4** | Myometrium diffusely heterogenous, asymmetrical wall thickening, with a focal area 38mm in the anterior wall that is isoechoic to the myometrium suggestive of adenomyosis with an adenomyoma | Irregular thickening of the junctional zone especially anteriorly and towards the fundus, junctional zone shows some cystic changes. Findings consistent with adenomyosis | * | Areas of increased stiffness throughout uterus |  |  |  |

The 3D MRE data acquired for one of the study participants diagnosed with adenomyosis (subject Ad2) are illustrated in comparison to corresponding data acquired for the control in Figure 2. The wave motion in x, y, and z directions (with corresponding zoomed in interpolated images) for the patient Ad2 (Figure 2 K (O), L (P), and M (Q)) has longer wavelength than for the control (Figure 2 B (F), C (G) and D (H)) indicating that regions of potential adenomyosis identified by the Radiologist, are potentially corresponding to higher stiffness values in the 3D MRE elastograms.

**Figure 2.**
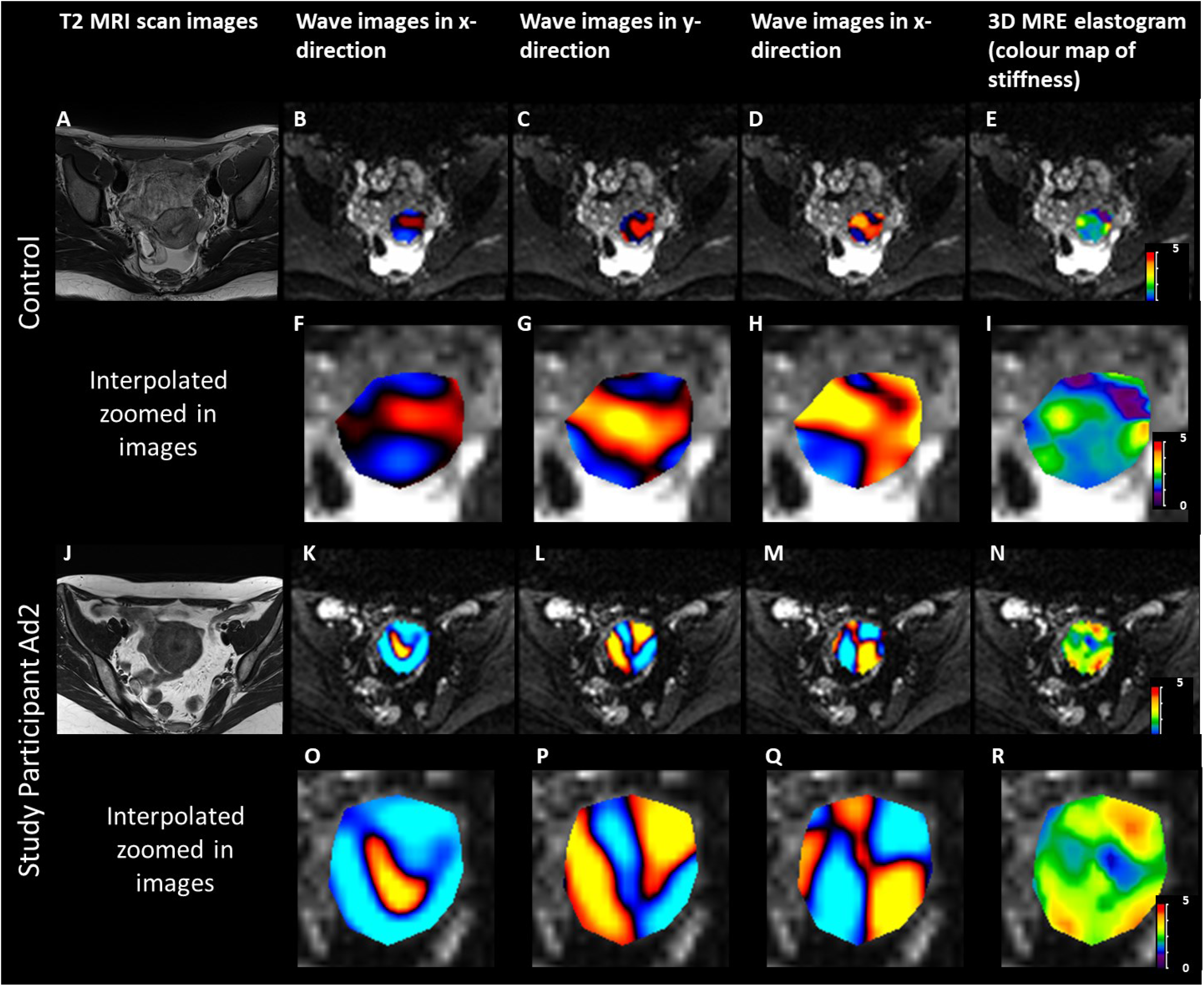
T2-weighted MR images, with 3D MRE waves images in x-, y- and z-directions and 3D MRE elastogram (colour map of stiffness) with a field of view of 24cm, superimposed on the MRE magnitude image of the uterus of a control subject and for the study participant Ad2 (scale bar 0 to 5 kPa). Corresponding zoomed in interpolated images with a field of view of 7.5cm. [to the reproduced in colour in print and on the web]

The measures of the stiffness of the whole uterus obtained for each participant are plotted in Figure 3. The median value obtained for the four patients with adenomyosis (2.93kPa; range 2.34 – 3.39kPa) is greater than the value measured for the healthy volunteer subject (2.04kPa).

**Figure 3.**
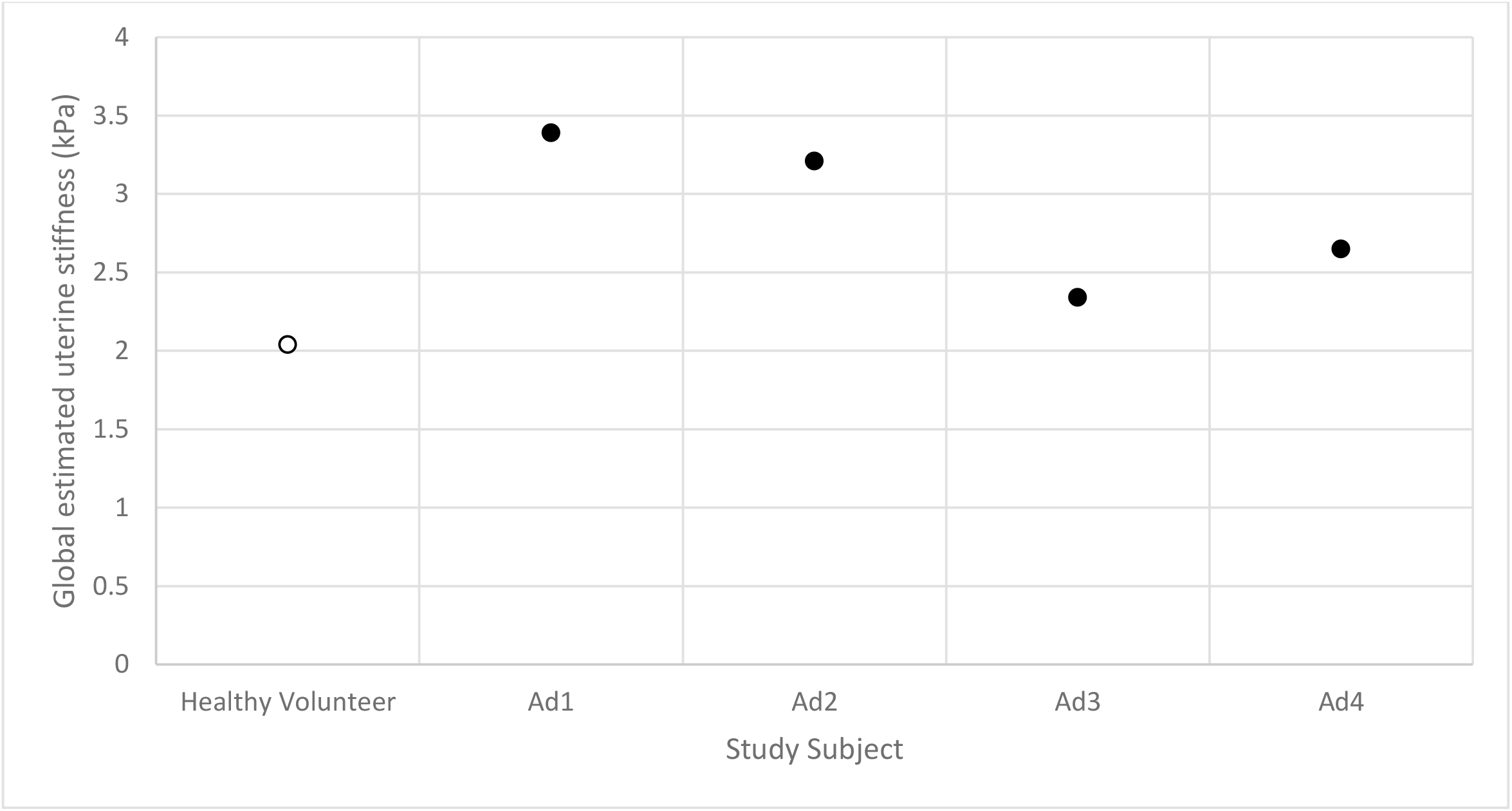
Average stiffness (kPa) of the whole uterus for the healthy volunteer subject and four patients (Ad1, Ad2, Ad3 and Ad4).

Figure 4 refers to the same patient as in Figure 2 (i.e., Ad2) and shows the resected uterus (A), the dissection (B) and sectioning that was performed so as to match with the T21 weighted MR images (C). The stained tissue sections depict the region where an adenomyotic lesion was detected during both the histopathological investigation of the uterus and on the T2 weighted MR images. The tissue has been systematically stained in (D), contains collagen fibres (E), smooth muscle fibres (F) and endometrial gland (G), consistent with the diagnosis of adenomyosis.

**Figure 4.**
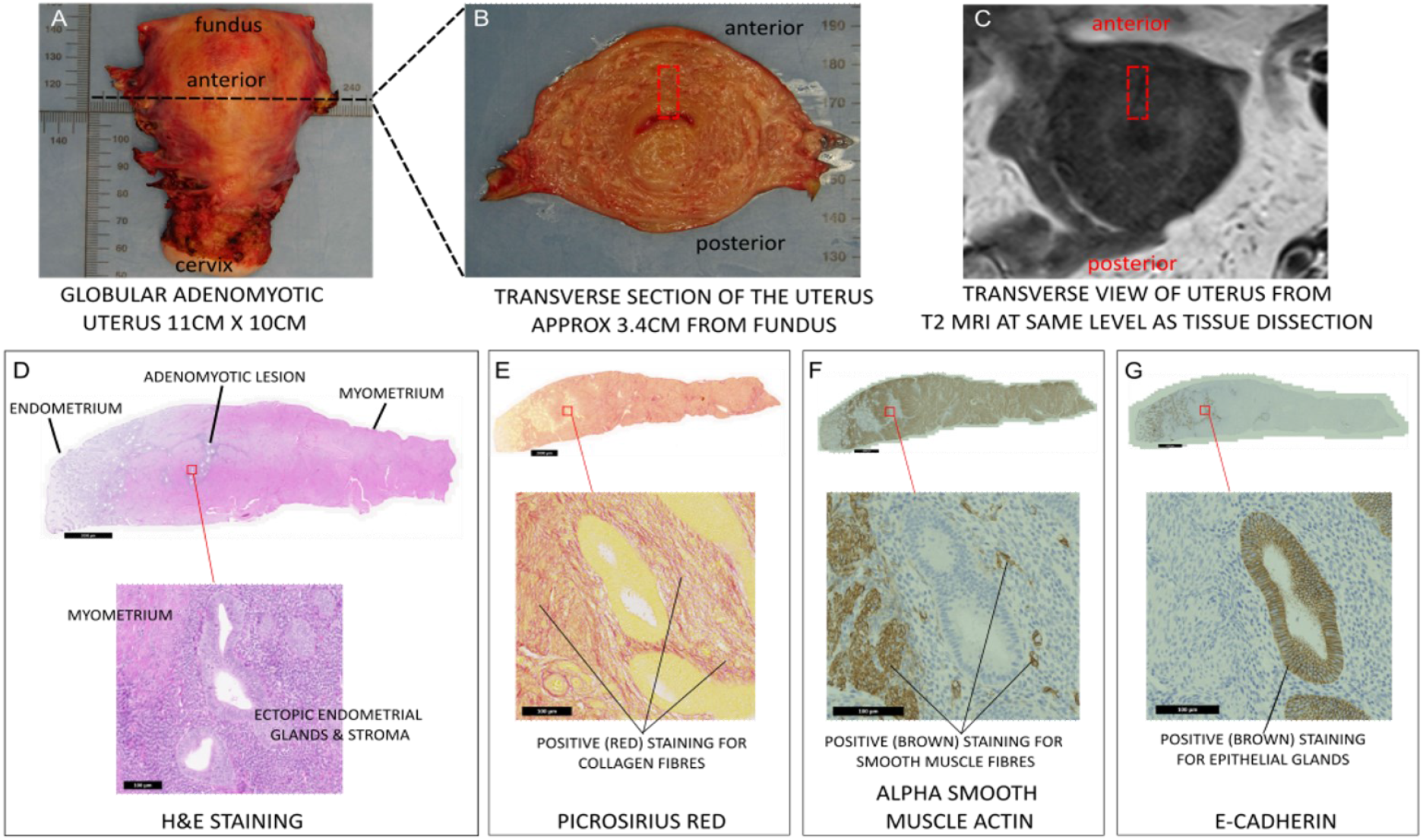
The whole uterus (A) was dissected in the transverse plane (B), in the direction and with section spacing that matched the series of T2 weighted MR images (C). The red rectangle depicts a region for which detailed histological analysis was performed and which confirmed the presence of endometrium, myometrium and an adenomyotic lesion (D), with ectopic endometrial glands and stroma of the adenomyotic lesion clearly visible in the magnified view. Picrosirius red staining indicated that excess collagen deposition was present (E), alpha smooth muscle actin staining confirmed the presence of smooth muscle fibres of myometrium (F), and e-cadherin staining provided support for the epithelial nature of the endometrial glands within the myometrial tissue (G). The scale bar corresponds to 2 mm. [to the reproduced in colour in print and on the web]

## Discussion

The results of this study provide preliminary evidence of the feasibility of applying MRE to evaluate global uterine stiffness in patients with suspected adenomyosis. The mean value of the stiffness of the whole uterus in the participants with adenomyosis was greater than that for the control subject, providing motivation for future studies with larger cohort sizes that would allow significance of this preliminary observation to be tested. Furthermore, for two participants in whom a hysterectomy was performed (for HMB), areas of suspected adenomyosis on T2-weighted MR images were confirmed histologically via stained uterine tissue sections, and mapped to corresponding elastograms obtained by using 3D MRE, suggesting increased tissue stiffness. These findings suggest that 3D MRE has the potential to allow tissue characterisation that is consistent with histopathology and which can be used to support non-invasive diagnosis of adenomyosis.

Further studies, which learn from this feasibility study, are needed to confirm these preliminary data and validate 3D MRE as a potential new non-invasive method of diagnosing and monitoring adenomyosis. The study of a larger cohort, which should include patients with and without diffuse adenomyosis, in the first instance, would allow for investigation into clinical utility of this diagnostic modality.

With regard to previous relevant research, Hobson et al (Hobson et al. 2007) performed ultrasound strain imaging on dissected uteri with confirmed adenomyosis and found no difference in the strain values obtained for the endometrium and myometrium, and concluded this was due to the pathological processes of the disease. However, more recently, in studies which used Transvaginal ultrasound elastography, Frank et al (Frank et al. 2016) have reported that adenomyotic lesions are softer than normal myometrium, whereas there have also been reports that regions of adenomyosis are stiffer than normal myometrium (Liu et al. 2018; Liu et al. 2016). Furthermore, Liu et al. (Liu et al. 2016) reported that the stiffness of regions of adenomyosis were greater in patients with heavier menstrual bleeding and this finding has been reproduced (Huang et al. 2022; Liu et al. 2018). Huang et al (Huang et al. 2022) also demonstrate a gradient in the density of markers of tissue stiffness within selected stained uterine tissue sections, from the region of adenomyosis (greater density) to the endometrium (less density). This possibly suggests the value of tissue stiffness as measured by elastography, may also change in relation to the proximity to the adenomyotic lesion. Transvaginal ultrasound elastography has also been used to demonstrate the differences between tissue stiffness for normal myometrium, uterine fibroids and regions of adenomyosis (Liu et al. 2018).

This suggests that the marker of tissue stiffness as measured by elastography, could be leveraged to non-invasively differentiate between uterine fibroids and adenomyosis, a clinically important differentiation to guide treatment for HMB.

One of the patients (Ad3) recruited to the present study was diagnosed as having focal adenomyosis, suggesting that some areas of the uterus appeared normal. This is consistent with the value of the stiffness of the uterus in this patient being closer to the value obtained for the control subject, rather than the other three adenomyosis patients (Ad1, 2, 4) who had histologically confirmed diffuse adenomyosis (see Figure 3).

The authors acknowledge there are limitations within this feasibility study. First, statistical analyses were restricted due to the small number of participants. Second, the MRE measurements obtained in the present study refer to the whole uterus and not to particular regions of adenomyosis identified by a Radiologist. The resolution of the 3D MRE is 3mm x 3mm x 3mm and therefore some of the histologically detected regions of adenomyosis could not be confirmed via T2 weighted MRI or 3D MRE as they were too small. This is the reason the tissue stiffness of the whole uterus, as opposed to regions of adenomyosis was utilised within this feasibility study. Despite these limitations, the present study does, however, have one major advantage. In previous MRE studies of the uterus, data have been acquired by using a 2D MRE technique (Stewart et al. 2011; Jondal et al. 2018; Obrzut et al. 2020), whereas a 3D EPI MRE sequence was used in the present study. To the best of our knowledge this is the first study where 3D MRE of the uterus has been accompanied with histological assessment from the same individual with adenomyosis.

Currently, there is no clear global consensus on the criteria for diagnosing adenomyosis by using either ultrasound or MRI techniques. The Morphological Uterus Sonographical Assessment (MUSA) provides a robust consensus on the definition of adenomyosis based on ultrasound imaging (Harmsen et al 2022). No such consensus exists for MRI diagnosis of adenomyosis. The data obtained from transvaginal ultrasound elastography is operator-dependant and the technique is not yet available in routine clinical practice. MRE, on the other hand, is a clinically approved diagnostic technique currently approved for use for grading of liver fibrosis and is widely available globally. The advantages of MRE over the other techniques are summarised in Table 2. Most importantly for clinical translation, MRE is not operator dependant. MRE is also truly non-invasive, compared to transvaginal ultrasound elastography which requires a transducer to be inserted into the vagina, and manual compression of the uterine tissues by means of the ultrasound transducer held by the operator, and may not be appropriate for use in younger patients and not acceptable to some women.

**Table 2.** The defining characteristics of current and possible future modalities that may be used for diagnosis of adenomyosis in comparison to the current gold standard of histopathological assessment of the myometrium.

|  | Hysterectomy<br>(histopathological<br>assessment of<br>myometrium) | Transvaginal<br>Ultrasound | Magnetic<br>Resonance<br>Imaging<br>(MRI) | Transvaginal<br>Ultrasound<br>Elastography | 3D Magnetic<br>Resonance<br>Elastography<br>(3D MRE) |
| --- | --- | --- | --- | --- | --- |
| Diagnosis accuracy<br>(if performed by<br>specialist) | 100% | ~83% | ~88% | To be<br>determined | To be<br>investigated |
| Estimated cost of<br>diagnosis | ++++ | + | ++ | Unknown | ++ |
| Non-invasive | No | No | Yes | No | Yes |
| Fertility sparing | No | Yes | Yes | Yes | Yes |
| Outpatient visit<br>(No hospital<br>admission required) | No | Yes | Yes | Yes | Yes |
| Quick | No | Yes | Yes | Yes | Yes |
| Monitor treatment<br>response | No | No | No | Unknown | Yes |
| Objective | Yes | No | Yes | No | Yes |
| Quantitative | No | No | No | No | Yes |

Global multicentre studies will be important to gather essential data to support the validation of these findings in both women with and without adenomyosis (no uterine pathology), before clinical translation and inclusion in the ongoing clinical health management of women with adenomyosis.

## Conclusion

The findings of the present study provide preliminary evidence of the feasibility of employing 3D MRE to aid in diagnosis of adenomyosis, a significant condition affecting women’s health.

## Data Availability

All data produced in the present study are available upon reasonable request to the authors.

## Acknowledgements

The authors would like to acknowledge: Ms. Catherine Murray for her help and support with patient recruitment; Professor Scott Semple, for his help and support with the MR investigations; Ms Moira Nicol, Dr Michael Millar and the team at SURF Histology, University of Edinburgh (https://surf.ed.ac.uk/facilities/histology/) for their advice and support with histological investigations; Dr. Bradley D. Bolster and Dr. Stephan Kannengiesser at Siemens Healthcare for their contribution to this work; all the patients and healthy volunteers who have generously donated their time, thoughts, tissue and blood samples, and much more to all of our studies. Funding: University of Edinburgh’s Institute for Regeneration & Repair (IRR) Early Career Researcher’s Innovation Award 2022, MRC Centre for Reproductive Health (MRC Centre grants: G1002033 and MR/N022556/1).

